# COVID-19 testing avoidance among patients with cardiovascular disease

**DOI:** 10.1101/2023.04.17.23288710

**Authors:** Koichiro Matsumura, Takahiro Tabuchi, Eijiro Yagi, Takeshi Ijichi, Misaki Hasegawa, Nobuhiro Yamada, Yohei Funauchi, Kazuyoshi Kakehi, Takayuki Kawamura, Gaku Nakazawa

## Abstract

**Background:** Rapid coronavirus 2019 (COVID-19) testing in symptomatic cases is extremely important for preventing the spread of COVID-19 infection and early therapeutic intervention. In contrast, whether symptomatic patients are tested depends largely on their health literacy, interpretation, and knowledge of COVID-19. We aimed to investigate the rate of COVID-19 testing avoidance despite having common cold symptoms in patients with cardiovascular disease and examine factors related to testing avoidance.

**Methods:** A large-scale epidemiological questionnaire survey, the Japan COVID-19 and Society Internet Survey 2022 (JACSIS), was conducted online from April to May 2022. The rate of COVID-19 testing avoidance was investigated in patients aged 20 to 80 years with cardiovascular risk factors (hypertension, dyslipidemia, or diabetes) or a history of cardiovascular disease (angina, myocardial infarction, or stroke), only those exhibiting common cold symptoms during the 2 months in the survey.

**Results:** Of the 1,565 eligible patients, 58% (909 patients) did not undergo COVID-19 testing. Multivariate analysis revealed that older age, obesity, non-walking regularly, long sedentary time, eating alone, frequent snacking, and having received 4 COVID-19 vaccinations were independently associated with testing avoidance.

**Conclusions:** In the chronic phase of the COVID-19 pandemic, prompt COVID-19 testing at the time of symptomatic disease is important, and strategies to reduce testing hesitancy should be considered.

## Introduction

The coronavirus disease of 2019 (COVID-19) pandemic poses challenges to health and welfare systems around the world. A combination of individual measures, including prevention of infection by masking and hand washing, early detection of infection by rapid COVID-19 testing, and isolation after infection, and social measures, including orders to stay home, social distancing, and avoidance of group activities, are used to control infection.^1,2^ Even with the availability of vaccines and effective treatments, efficient identification and control of infection is critical and needs to continue. In particular, surveillance is a core function of the public health system and is essential for the control of COVID-19.^3^ Japan’s surveillance strategy is based on testing of symptomatic individuals in the community and supports an effective public health response to COVID-19.^4,5^ However, the strength of this strategy depends on adequate inspection practices in the community. Even if testing is convenient and public health advisories indicate the importance of testing, the general population may still not be adequately compliant. A number of social factors, such as a less symptomatic patient population, accessibility issues to testing sites, isolation, and the need to take time off work, promote testing hesitancy.^6^ Decreased test-taking behaviors in the population risk amplification of infection. Therefore, effective surveillance is essential for the detection and control of COVID-19, especially in patients with chronic disease, including cardiovascular disease, as these individuals are at high risk. Having a system in place to detect infections as early as possible is important, both when the number of infections is high and when the community is largely under control.^7^ While many studies focusing on vaccine hesitancy have been conducted, research on COVID-19 test hesitancy is limited. Therefore, we utilized a large epidemiological database obtained from an online survey to analyze the hesitancy of COVID-19 testing in patients with cardiovascular disease.

## Methods

### Study Design

Data from the Japan COVID-19 and Society Internet Survey 2022 (JACSIS 2022) were obtained from 32,000 participants aged 15–80 during the period of April 1 to May 30, 2022 (6th wave) (Figure 1). The survey data were collected online using a self-reported questionnaire designed to investigate changes in social issues such as health, medical care, work styles, and the economy during the COVID-19 pandemic. The survey was conducted by a Japanese Internet research company (Rakuten Insight, Inc., Tokyo, Japan) comprising approximately 2.2 million panelists from diverse socioeconomic backgrounds on a national scale. Data will be shared on request to the corresponding author.

**Figure 1.**
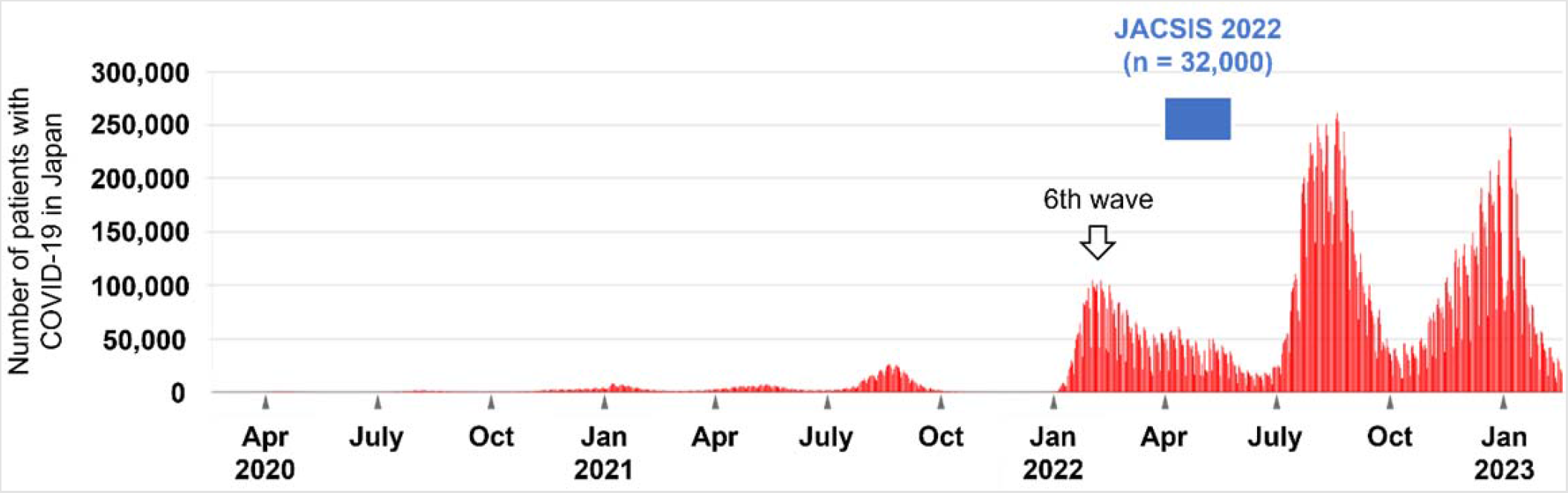
Investigation periods of JACSIS 2022 which conducted between April 1 and May 30, 2022 (during 6th wave in Japan). Abbreviations: COVID-19, coronavirus disease of 2019; JACSIS, Japan COVID-19 and Society Internet Survey.

### Target population and outcome

In patients aged 20 to 80 years with cardiovascular risk factors (hypertension, dyslipidemia, or diabetes) or a history of cardiovascular disease (angina, myocardial infarction, or stroke), only those exhibiting common cold symptoms were extracted from the JACSIS 2022 data base, during the 2 months in which the survey was conducted. The rate of COVID-19 testing avoidance was investigated in these patients.

### Managing data quality

Participants were recruited for the JACSIS 2022 using a random sampling method generated by a computer algorithm, according to the official Japanese demographic structure based on age, gender, and prefecture of residence. To verify the data quality, respondents who provided inconsistent or artificial/unnatural responses were excluded. To detect such discrepancies, the following 3 directives were used: “Please choose the second one from the bottom,” “Please answer affirmatively to all questions about drug use,” and “Please answer affirmatively to all questions about chronic disease status.”

### Covariates

For clinical background factors, we extracted data on age, sex, body mass index, history of cancer, history of depression, smoking status, and alcohol consumption. For socioeconomic factors, we analyzed data on type of residence, marital status, the presence of family members living with the patient, and employment status. The following data were collected regarding the patients’ daily attitudes and lifestyle: frequency of teleworking, duration of walking, sedentary time, sleeping time, frequency of eating alone, missing breakfast, frequency of snacking, duration of smartphone use, and number of COVID-19 vaccinations. Patients were grouped according to the following definitions: working at least 1 day per week without going to the workplace was defined as “teleworking”; walking an average of at least 1 hour per day was defined as “walking regularly”; sitting an average of at least 6 hours per day as “long sedentary time”; sleeping fewer than 6 hours per day on average as “short sleep”; eating meals alone at least twice a week as “eating alone”; eating breakfast regularly as “having breakfast regularly”; eating snacks more than twice per week as “frequent snacking”; and using a smartphone for more than 4 hours a day on average as “prolonged smartphone use”.

To investigate whether loneliness and fear of COVID-19 affected COVID-19 testing avoidance, loneliness was assessed using the 3-item University of California, Los Angeles (UCLA) loneliness scale, and fear of COVID-19 was assessed using the Japanese version of theFCV-19S.^8,9^ The 3-item UCLA loneliness score ranged from 3 to 12, with a higher score indicating more severe loneliness. Based on previous reports, 6 or higher was defined as loneliness.^8,10^ The FCV-19S asked respondents to indicate their degree of agreement with each question on a scale of 1 (strongly disagree) to 5 (strongly agree) points. The FCV-19S consisted of 7 questions, with total scores ranging from 7 to 35 points: Question 1 “I am most afraid of COVID-19”, Question 2 “It makes me uncomfortable to think about COVID-19”, Question 3 “Sweaty hands when thinking about COVID-19”, Question 4 “Afraid of losing my life because of COVID-19”, Question 5 “I get nervous and anxious when watching news and topics about COVID-19 on social media”, Question 6 “I cannot sleep worrying about getting infected with COVID-19”, and Question 7 “Thinking about COVID-19 causes rapid heartbeat and palpitation”. Based on previous reports, a score of 21 or higher was defined as having a fear of COVID-19.^9^

### Questionnaire on sources of medically relevant information and reasons for avoidance of testing

Questions about sources of healthcare-related information regarding COVID-19 infections were asked in a multiple-response format for the following items: family or friends, workspace, healthcare providers, government offices, on the Internet, and on television. Patients with COVID-19 testing avoidance were asked to select from the following 7 reasons in a multiple-answer format: I did not see anyone around me who was infected; I did not think I was infected; I tried to get an exam but could not make an appointment; I did not trust the accuracy of the tests; It is a hassle to communicate with the health department if you test positive; I would have to miss work if I tested positive; and I was afraid of testing positive and having my family and workplace find out.

### Statistical analysis

Categorical variables are presented as numbers and percentages. Differences between groups were analyzed using the chi-square test. Multivariate logistic regression analysis was performed to examine the factors related to the absence of COVID-19 testing. Statistical significance was set at a p value < 0.05. All statistical analyses were performed using JMP 16.0.0 (SAS Institute Inc., Cary, NC, USA).

### Ethics

This study adhered to the principles outlined in the Declaration of Helsinki and the protocol was approved by the Ethics Committee of the Osaka International Cancer Institute (approval number: 20084-6). All provided informed consent online when enrolling for this survey.

## Results

### Patient characteristics

Based on our inclusion criteria, 1,556 patients with cardiovascular disease were selected from the JACSIS 2022 database (mean age, 53 years; 42% female). Within 2 months of the survey, 58% (909 of 1,556 patients) had not undergone COVID-19 testing despite having common cold symptoms (Figure 2A).

**Figure 2.**
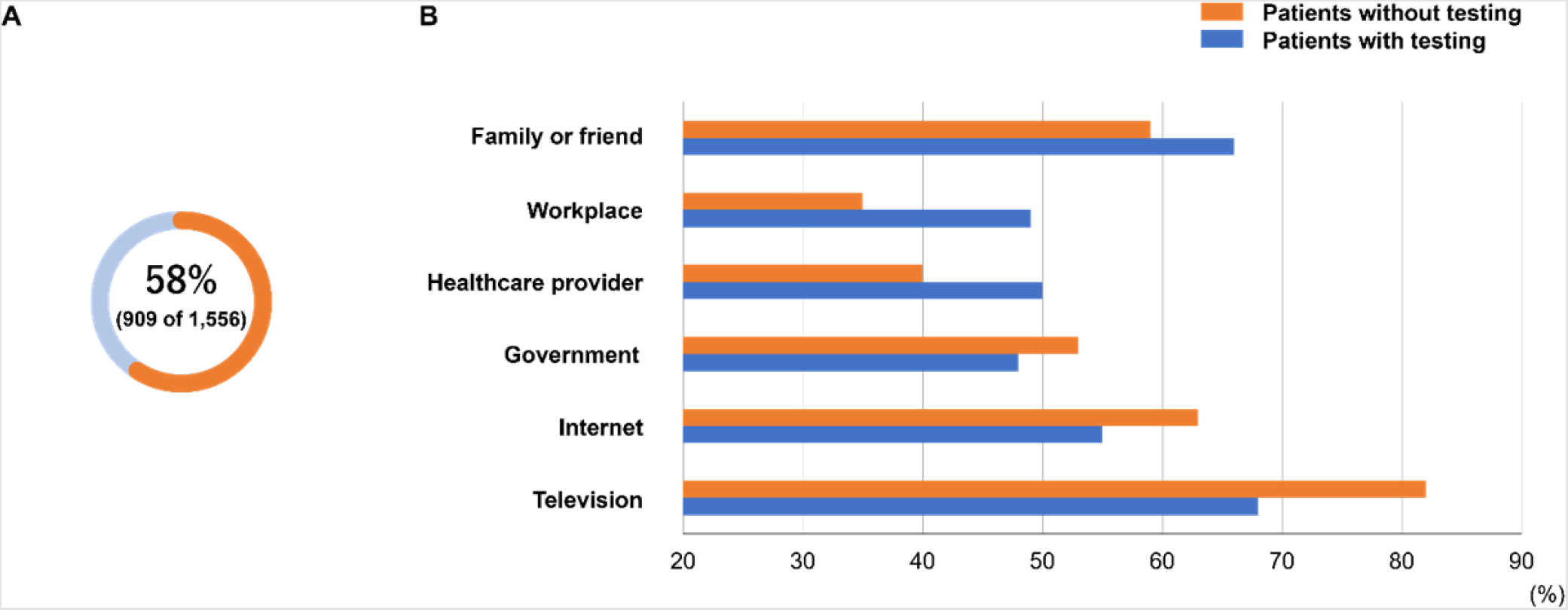
Rate of COVID-19 testing avoidance in patients with cardiovascular disease (A) and self-reported sources of available medically relevant information on COVID-19 (B) Abbreviations: COVID-19, coronavirus disease of 2019.

Patients who did not take a test were significantly more likely to be elderly, female, obese, nonsmokers, homeowners, and unemployed than those who did test (Table 1). However, the proportions of smoking, alcohol use, marital status, and living alone were similar between the two groups. Those who were not tested were less likely to do teleworking and walked more regularly than those who were (Table 2). Those who did not test were more likely to have long sedentary time, eat alone, have breakfast regularly, and consume frequent snacks than those who did test. Approximately 80% (1,212 of 1,556) of patients in both groups had received 3 or more doses of the COVID-19 vaccine.

**Table 1.**
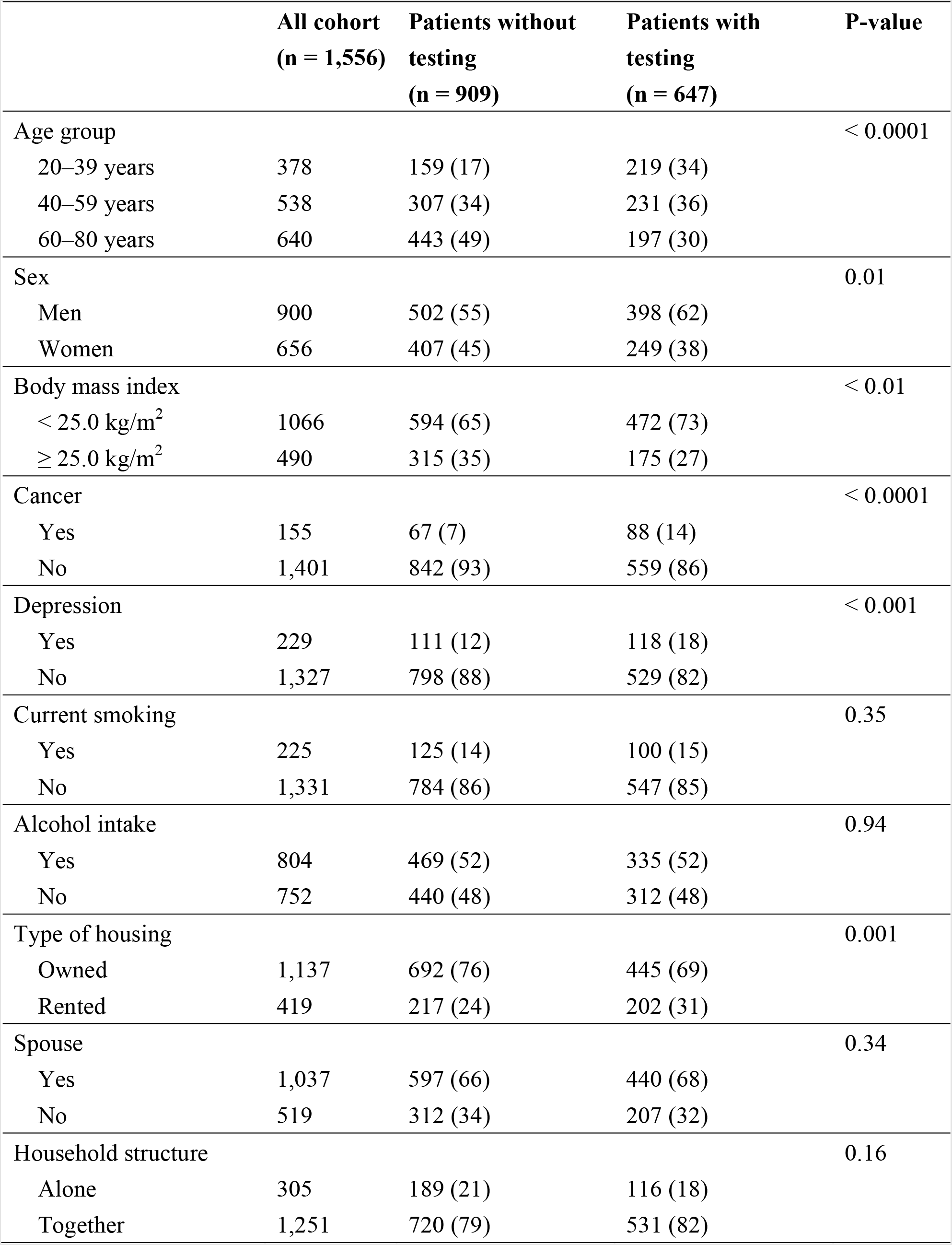
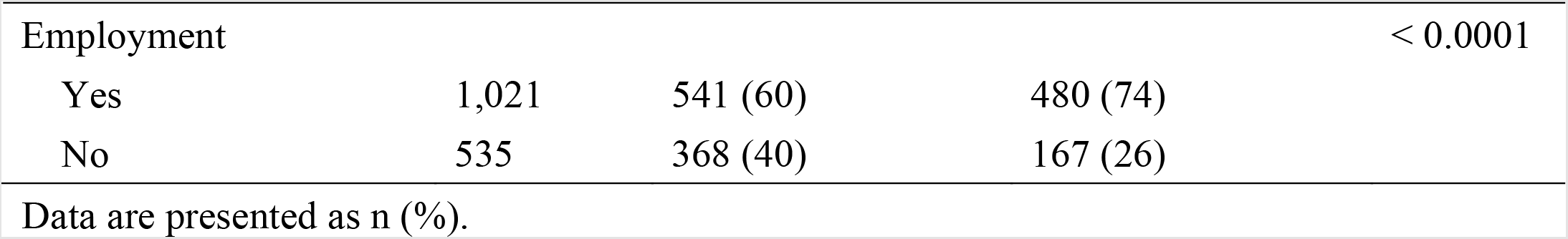
Patient characteristics

|  | **All cohort**<br>**(n = 1,556)** | **Patients without**<br>**testing**<br>**(n = 909)** | **Patients with**<br>**testing**<br>**(n = 647)** | **P-value** |
| --- | --- | --- | --- | --- |
| Age group |  |  |  | < 0.0001 |
| 20–39 years | 378 | 159 (17) | 219 (34) |  |
| 40–59 years | 538 | 307 (34) | 231 (36) |  |
| 60–80 years | 640 | 443 (49) | 197 (30) |  |
| Sex |  |  |  | 0.01 |
| Men | 900 | 502 (55) | 398 (62) |  |
| Women | 656 | 407 (45) | 249 (38) |  |
| Body mass index |  |  |  | < 0.01 |
| < 25.0 kg/m<sup>2</sup> | 1066 | 594 (65) | 472 (73) |  |
| ≥ 25.0 kg/m<sup>2</sup> | 490 | 315 (35) | 175 (27) |  |
| Cancer |  |  |  | < 0.0001 |
| Yes | 155 | 67 (7) | 88 (14) |  |
| No | 1,401 | 842 (93) | 559 (86) |  |
| Depression |  |  |  | < 0.001 |
| Yes | 229 | 111 (12) | 118 (18) |  |
| No | 1,327 | 798 (88) | 529 (82) |  |
| Current smoking |  |  |  | 0.35 |
| Yes | 225 | 125 (14) | 100 (15) |  |
| No | 1,331 | 784 (86) | 547 (85) |  |
| Alcohol intake |  |  |  | 0.94 |
| Yes | 804 | 469 (52) | 335 (52) |  |
| No | 752 | 440 (48) | 312 (48) |  |
| Type of housing |  |  |  | 0.001 |
| Owned | 1,137 | 692 (76) | 445 (69) |  |
| Rented | 419 | 217 (24) | 202 (31) |  |
| Spouse |  |  |  | 0.34 |
| Yes | 1,037 | 597 (66) | 440 (68) |  |
| No | 519 | 312 (34) | 207 (32) |  |
| Household structure |  |  |  | 0.16 |
| Alone | 305 | 189 (21) | 116 (18) |  |
| Together | 1,251 | 720 (79) | 531 (82) |  |
| Employment |  |  |  | < 0.0001 |
| Yes | 1,021 | 541 (60) | 480 (74) |  |
| No | 535 | 368 (40) | 167 (26) |  |

Data are presented as n (%).

**Table 2.**
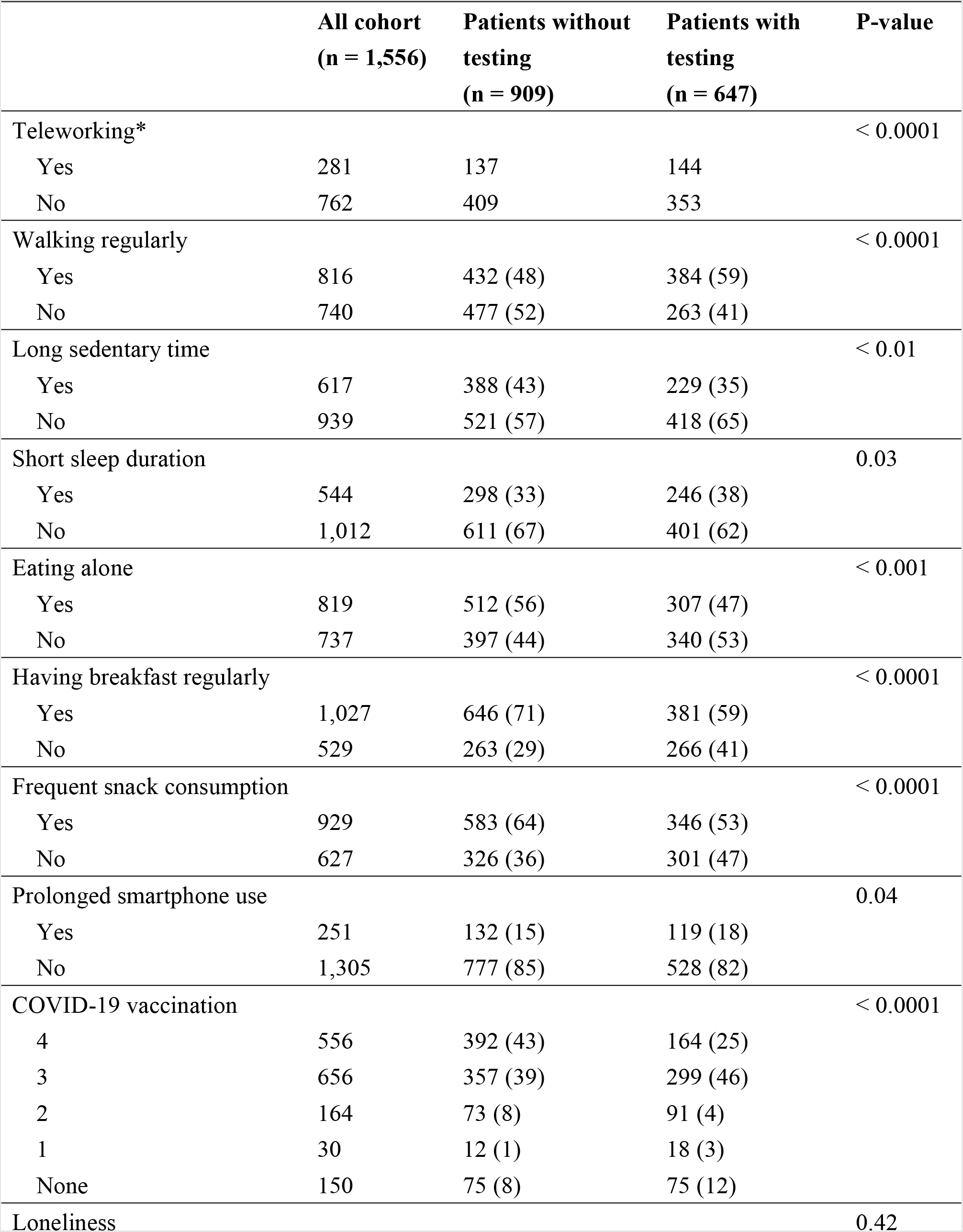
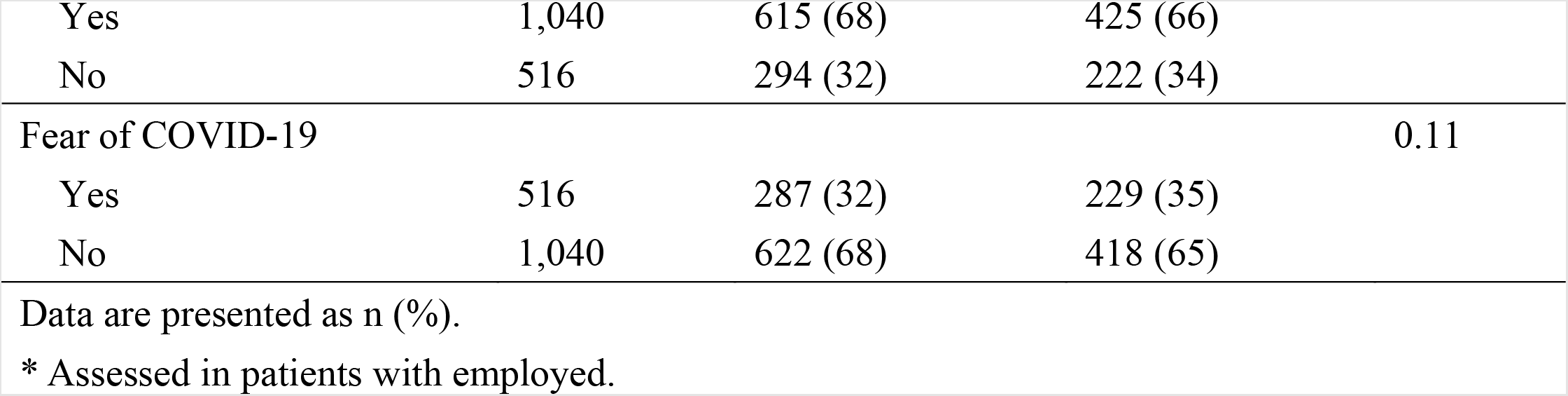
Lifestyle characteristics

### Multivariate analysis

A multivariate logistic regression analysis was performed to examine the factors associated with COVID-19 test avoidance (Table 3). Compared to younger patients (20-39 years), middle-aged to older patients were positively associated with test avoidance. Obesity (body mass index ≥ 25.0 kg/m^2^), long sedentary time, eating alone, and frequent snack consumption were positively associated with test avoidance. Having received 4 COVID-19 vaccinations was also positively associated with testing avoidance, compared to those who had received 3 or less COVID-19 vaccination. Regular walking was negatively associated with test avoidance.

**Table 3.** Multivariate logistic regression analyses of factors associated with testing hesitancy

|  | **OR (95% CI)** | **P value** |
| --- | --- | --- |
| Age group |  |  |
| 20–39 years | Reference |  |
| 40–59 years | 1.45 (1.07–1.95) | 0.02 |
| 60–80 years | 1.74 (1.20–2.50) | < 0.01 |
| Female sex | 1.04 (0.81–1.32) | 0.76 |
| Body mass index $\geq 25.0$ kg/m<sup>2</sup> | 1.36 (1.06–1.73) | 0.01 |
| Cancer | 0.85 (0.57–1.27) | 0.44 |
| Depression | 0.91 (0.65–1.28) | 0.60 |
| Current smoking | 1.11 (0.81–1.51) | 0.52 |
| Alcohol intake | 0.91 (0.73–1.14) | 0.43 |
| Rented housing | 0.84 (0.64–1.10) | 0.21 |
| Presence of spouse | 0.79 (0.59–1.07) | 0.13 |
| Living alone | 1.25 (0.87–1.79) | 0.22 |
| Unemployed | 0.47 (0.16–1.35) | 0.16 |
| Teleworking | 1.08 (0.79–1.47) | 0.62 |
| Walking regularly | 0.74 (0.59–0.92) | < 0.01 |
| Long sedentary time | 1.31 (1.04–1.65) | 0.02 |
| Short sleep duration | 1.03 (0.81–1.30) | 0.81 |
| Eating alone | 1.28 (1.02–1.63) | 0.03 |
| Having breakfast regularly | 1.20 (0.93–1.54) | 0.17 |
| Frequent snack consumption | 1.31 (1.04–1.64) | 0.02 |
| Prolonged smartphone use | 0.96 (0.71–1.30) | 0.79 |
| COVID-19 vaccination |  |  |
| 4 | Reference |  |
| 3 | 0.62 (0.47–0.83) | 0.001 |
| 2 | 0.52 (0.34–0.79) | 0.001 |
| 1 | 0.52 (0.23–1.19) | 0.12 |
| None | 0.58 (0.38–0.89) | 0.01 |
| Loneliness | 1.21 (0.95–1.53) | 0.12 |
| Fear of COVID-19 | 0.88 (0.70–1.11) | 0.28 |

Abbreviations: CI, confidence interval; COVID-19, coronavirus disease 2019; OR, odds ratio Adjusted for all listed variables.

### Reasons for not having been tested for COVID-19

A total of 462 patients responded to the questionnaire regarding why they had not tested (Table 4). Many patients did not believe that they were infected and did not undergo testing because they did not know anyone around them who was infected. Additionally, a few feared that others would find out when they tested positive.

**Table 4.** Reasons for not testing for COVID-19

| **Questions** | **Number of patients**<br>**(n = 462)** |
| --- | --- |
| I did not see anyone around me who was infected. | 159 (36) |
| I did not think I was infected. | 145 (32) |
| I tried to get an exam but could not make an appointment. | 14 (3) |
| I did not trust the accuracy of the tests. | 13 (3) |
| It is a hassle to communicate with the health department if you test positive. | 10 (2) |
| I would have to miss work if I tested positive. | 10 (2) |
| I was afraid of testing positive and having my family and workplace find out. | 5 (1) |

Data are presented as n (%).

### Sources of medically relevant information on COVID-19

Sources of healthcare-related information on COVID-19 were as follows for patients without testing vs. patients with testing: family or friend, 59% (536/909) vs. 66% (427/647), p < 0.01; workspace, 35% (321/909) vs. 49% (314/647), p < 0.0001; healthcare provider, 40% (364/909) vs. 50% (324/647), p < 0.0001; government, 53% (481/909) vs. 48% (309/647), p = 0.04; Internet, 63% (574/909) vs. 55% (355/647), p = 0.001; television, 82% (748/909) vs. 68% (438/647), p < 0.0001 (Figure 2B). The overall trend was that more patients obtained information from television, followed by family or friends and the Internet.

## Discussion

During the early stages of the COVID-19 pandemic in 2020, the establishment of testing systems was delayed, and adequate testing could not be conducted. In addition, COVID-19 was not well understood, and various social and psychological barriers such as stigma, misinformation, low health literacy, and low awareness led to hesitancy in testing.^6^ As the COVID-19 pandemic enters the chronic phase, testing systems have been established, COVID-19 is better understood, and barriers to testing have decreased. For these testing systems to be effective in preventing the spread of infection, it is important that a series of steps be taken, including accurate diagnosis of infected persons through adequate testing, isolation of infected patients, and notification of close contacts. The COVID-19 testing rate in Japan is high, even according to international standards, and public health advisories strongly emphasize the importance of testing, even for minor symptoms. In this study, however, we found that approximately 60% of patients with cardiovascular disease were not tested despite having common cold symptoms. This result is not necessarily surprising as survey of community residents in other countries collected from 2020 to 2021 also reported a high rate of failure to test when symptomatic.^7,11,12^ Social strategies to prevent infection have a spectrum of negative effects on economic, occupational, health, and lifestyle parameters. Thus, health literacy declines significantly over time.^13,14^ Declining health literacy due to prolonged pandemics is a threat to the effectiveness of public health strategies to control COVID-19. The results of this study, which examined the actual situation in the chronic phase of the pandemic, suggest the need to re-examine the social strategies of COVID-19 testing.

An overabundance of valid and invalid information related to the COVID-19 pandemic was spread via digital media, a phenomenon known as infodemic.^15^ Health literacy is the ability to correctly understand, evaluate, and apply medical information, which is important to address the infodemic of COVID-19. A cross-sectional survey on COVID-19 health literacy found that approximately 50% of the participants had difficulty determining whether they could trust media coverage of COVID-19.^13^ People with low health literacy tend to have less of an understanding of COVID-19 symptoms and are less willing to fully implement actions to prevent infection.^14^ In addition, individuals with lower health literacy are less likely to view social distancing as important in preventing infection and are more likely to perceive themselves as less likely to be infected.^16^ Socially vulnerable and older adults tend to have lower health literacy, suggesting the need to address COVID-19-related health disparities in these populations.^17^ A cross-sectional telephone survey conducted in Chicago at the beginning of the 2020 pandemic reported that among patients with chronic conditions, those with low health literacy were more likely to believe they would not be infected with COVID-19.^18^ Furthermore, approximately 25% of those surveyed said they did not believe they had any chance of contracting COVID-19. These patients tended to trust information from government agencies. Self-affirmation is a central concept in shaping health literacy.^19^ Low self-affirmation, low health literacy, and a socioeconomically disadvantaged status were associated with COVID-19 test hesitancy.^14^

In a survey conducted by Dayton et al. during 2020–2021, the most common reason for not getting tested was that close contacts were informed of the test results.^20^ Furthermore, in their survey, individuals who were unvaccinated were more hesitant to be tested. However, in our study, few patients indicated that their reason for not testing was fear that their families or workplaces would find out. The reasons for not testing may be the result of changing stigma and social values between the early and chronic phases of the pandemic. In fact, patients with 4 vaccinations avoided testing more frequently than those who were less than 4 vaccinated, indicating that being fully vaccinated may provide reassurance against the risk of infection.

Sharing COVID-19 information with family and friends has been reported to influence individual health literacy and prevention behaviors.^21^ Health literacy is not just an individual skill, but also a distributed resource available within social networks.^14^ In addition to interventions to improve individual health literacy, strategies to enhance information sharing and support within close social networks may be particularly useful for vulnerable populations with low health literacy who rely on fewer sources of information. Such strategies have the potential to reduce social disparities in health literacy while leading improvements in risk communication and creating behavioral changes at a time of heightened health risk.

### Strengths and limitations

Although many studies have been conducted on COVID-19 vaccine avoidance, there have been few reports on COVID-19 testing hesitancy. Previous reports on COVID-19 testing avoidance have mainly focused on social factors, such as laboratory center hours, inaccessibility, location of centers, commercial strategies, and decisions on how to assign tests.^6,7,11,12^ In addition, several studies on stigma in the early stages of COVID-19 pandemic have been conducted.^20,22^ Our study focused on patient attributes related to COVID-19 test avoidance, including clinical factors and socioeconomic background. Our combined analysis of lifestyle and medical information sources is particularly instructive. In addition, social policies for infection prevention have decreased significantly over time.^13,14^ Therefore, we believe that our study of COVID-19 testing avoidance in the chronic phase of the pandemic provides valuable clinical, social, and public health information. Additionally, the results of this study will significantly contribute to future societal policies regarding COVID-19 in the cardiovascular field.

This study had several limitations. First, the use of a questionnaire is invariably accompanied by sampling, selection, and response biases. However, a computer algorithm used a random sampling method according to age group, gender, and area of residence in Japan in an effort to obtain robust data with minimal bias. In addition, inconsistent, artificial, or unnatural responses were excluded from the sample to ensure quality. Second, because the survey was self-reported, the responses reflected the participants’ own perceptions, which may have differed from objective abilities and behaviors. Further research using objective measures is required to confirm the relationships suggested in this study.

## Conclusions

The decline in health literacy due to the prolonged pandemic has resulted in the avoidance of COVID-19 testing in many patients with cardiovascular disease, indicating a need to reconsider and address the social systems around COVID-19 testing.

## Data Availability

Data will be shared on request to the corresponding author.

## Non-standard Abbreviations and Acronyms

JACSIS: the Japan COVID-19 and Society Internet Survey 2022

## Acknowledgments

We would like to thank Editage (www.editage.com) for English language editing.

